# Effect modification in network meta-analyses of treatments for relapsing refractory multiple myeloma (RRMM): systematic review, meta-analysis, and simulation

**DOI:** 10.1101/2022.05.13.22275047

**Authors:** Christopher James Rose, Ingrid Kristine Ohm, Liv Giske, Gunn Eva Næss, Atle Fretheim

## Abstract

**Aims:** Network meta-analysis (NMA) has been used in several systematic reviews on relapsing refractory multiple myeloma (RRMM). NMAs have been questioned on the basis that effect modification may invalidate the underpinning assumptions. We aimed to systematically review and meta-analyze the evidence for effect modification of hazard ratios (HRs) for overall survival (OS) and progression-free survival (PFS) with respect to refractory status and number of treatment lines.

**Methods:** We extracted stratified HR estimates from 42 phase 2 and 3 randomized controlled trials (RCTs). We tested for within-study effect modification and used meta-analyses to estimate ratios of hazard ratios (RHRs) across trial under assumptions that strongly favor the modification hypothesis. RHR estimates were used in simulations to estimate how many NMA results would be expected to differ in the presence versus absence of effect modification.

**Results:** Most (95%) publications could have reported stratified estimates but only 14% (OS) and 43% (PFS) did. Within-study evidence for effect modification is very weak (*p* > 0.05 for 47 of 49 sets of stratified estimates). The largest RHR estimated was 1.31 (95% CI 1.16–1.47), for the modifying effect of refractory status on HR for PFS. Simulations suggest that, in the worst case, effect modification would result in 4.48% (95% CI 4.42%–4.53%) of NMA estimates differing statistically significantly in the presence versus absence of effect modification.

**Conclusions:** Effect modification is essentially undetectable in phase 2 and 3 trials. In the worst case, it is unlikely to affect more than about 5% of random-effects NMA estimates.

## INTRODUCTION

A defining characteristic of relapsing refractory multiple myeloma (RRMM) is that patients either do not respond, or stop responding — i.e., are refractory — to specific treatments.^1^ Refractory patients must switch to alternative treatments, if available. Multiple treatment options now exist, and the treatment regimens often comprise multiple drugs in combination. This naturally leads to questions about treatment superiority. These have been addressed in several systematic reviews that have used network meta-analysis (NMA).^2,3,4,5,6,7^

If the assumptions underpinning an NMA model are sufficiently satisfied, NMA facilitates meta-analytical estimation of all pairs of treatment effects, including between treatments that have not been compared directly in a trial. One of these assumptions is called the transitivity assumption^8,9,10^ which, informally, means that a treatment effect for one comparison can be calculated by adding or subtracting treatment effects for other comparisons in the network. This allows treatment effects to be estimated for pairs of treatments that have not been directly compared by the trials included in the network (i.e., indirect comparisons).

NMAs should assess and report on the likely validity of the transitivity assumption. This requires comparing distributions of effect modifiers across trials.^8^ An effect modifier is a variable that causes a difference in treatment effect but is not itself a treatment or an outcome.^11,12^ In plain English: effect modification is about stratification — when effect modification occurs, treatment effect is different for different subgroups of patients. It is important to distinguish between a variable that is associated with *treatment effect* (a comparison between treatments) and a variable that is only associated with *outcome* (e.g., overall survival for a particular patient). The former is an effect modifier, but the latter is a risk factor. Number of lines of treatment (LOT) is presumably a risk factor for overall survival, if for no other reason than patients who have received many LOT will be older. However, that does not mean it is also an effect modifier.

Unfortunately, non-statistical articles on NMA often conflate risk factors and effect modifiers when considering the transitivity assumption. Risk factors are not a concern for NMAs of RCTs because, in expectation, randomization excludes the possibility that they account for observed treatment effects. In large part, this is why RCTs are so useful. However, if a *fixed-effects* NMA is applied to estimates from trials with different distributions of effect modifiers, the transitivity assumption will be threatened because the estimates have different interpretations, and with it, the validity of the NMA. That said, the nature and extent to which an NMA may be invalidated by effect modification depends on the magnitudes and directions of the modifications. If modification is small compared to the precisions of the trial estimates, NMA estimates may still be consistent with the true treatment effects (e.g., confidence intervals may contain the target parameter values). *Random-effects* NMAs are designed specifically to address heterogeneity in trial-level treatment effects.

The use of NMA in RRMM has been criticized^13^ on the basis that variables such as refractory status and LOT are effect modifiers, with the implication that NMAsthat do not account for effect modification may be untrustworthy. The present article was motivated by an ongoing health technology assessment (HTA) we are conducting on treatments for RRMM that was commissioned via Norway’s National System for Managed Introduction of New Health Technologies within the Specialist Health Service (“Nye Metoder”).^14^ One of our clinical advisors highlighted concerns about effect modification with respect to refractory status and LOT. While these concerns have been raised in previous work,^13^ we could not find definitive quantitative research on effect modification in RRMM that could inform our HTA. We therefore performed a systematic review and meta-analysis of stratified estimates reported by the trials included in our HTA. We then used the meta-analysis results in a simulation study to assess the degree to which NMA estimates are likely to be affected by effect modification.

## METHODS

This meta-analysis was not prespecified or registered because it was performed in response to comments on an ongoing HTA. Further methodological details and completed PRISMA checklists^15^ are available in Supplementary Materials.

### Literature search strategy

The search was first performed in February 2020 and was regularly updated until January 2022 (ongoing trials until June 2021). We limited the search to RCTs, used the search term Multiple Myeloma, and used MeSH-terms and text words. Halfway through we limited the search to include the terms Relapse or Refractory. The full strategy is presented in Supplementary Materials. We also contacted project stakeholders, including industry, to solicit suggestions for potentially relevant publications. We did not systematically search beyond this work to support our HTA because we are primarily interested in effect modification within the trials included in our HTA. Via manual searching we found nine articles reporting stratified estimates for the included trials^16,17,18,19,20,21,22,23,24^ but used stratified estimates from the main trial publications, because they are more likely to have been prespecified.

### Inclusion and exclusion criteria

From the identified publications, we included those that provide estimates of hazard ratios (HRs) for OS or PFS that could be included in NMAs (i.e., those that report point estimates and a statement of precision such as a confidence interval or *p*-value). We excluded trials comparing doses or schedules of the same treatment.

We excluded publications from meta-analysis if they did not report stratified estimates of HR for all strata for at least one of two potential effect-modifiers (e.g., we would have excluded a study if it did report an estimate for lenalidomide-refractory patients but did not report an estimate for patients not refractory to lenalidomide). We excluded publications that did not report numerical statements of uncertainty on stratified estimates (e.g., we excluded one study that reported point estimates numerically but only provided a graphical presentation of the confidence intervals).

### Data extraction

CJR extracted data from trial reports and supplementary materials. Extracted data were checked independently by IKO. Disagreements were resolved via discussion. We extracted estimates of HR for OS and PFS, stratified by LOT and refractory status or previous use of immunomodulatory drugs (IMiDs; see below), where available. For LOT, we extracted data using the categorizations used by the publications (e.g., Attal 2019 used 2–3 vs >3 LOT, while Dimopoulos 2016 used 1 vs 2 vs 3 vs >3 LOT). For refractory status we preferentially extracted estimates stratified by refractory status with respect to lenalidomide (Revlimid), which was identified as being of particular concern by our clinical advisor (i.e., if stratified estimates were available for lenalidomide-refractory patients and patients not refractory to lenalidomide, we extracted these estimates). If this information was not reported, we extracted estimates for refractory status with respect to other named IMiDs such as thalidomide (of which lenalidomide is an analog). If this information was not reported, we extracted estimates for refractory status with respect to IMiDs in general. If this information was not available, we extracted estimates stratified by previous use of lenalidomide or other IMiDs, on the assumption that previously IMiD use is a reasonable proxy for being refractory to an IMiD. For the same reason, we did not extract estimates stratified by LOT and refractory status simultaneously (e.g., comparing OS in lenalidomide-refractory patients with one LOT). In summary, we used a pragmatic and inclusive definition of refractory status rather than a strict definition that would have yielded very little evidence on effect modification. For brevity we call this concept “refractory status” in the remainder of the article. However, readers are reminded that the concept is broader than this name implies (the statistical analysis section describes how we address the issue statistically).

### Statistical analyses

We first performed pairwise random-effects meta-analyses of stratified HRs, grouped by trial, for refractory status and LOT. This facilitates testing for evidence of effect modification within each trial. Because these analyses yielded very weak evidence for effect modification, but there nevertheless seems to be strong opinions that effect modification does occur and is a problem for NMAs for RRMM, we then performed pairwise random-effects meta-analyses of ratios of hazard ratios (RHRs; described below) for refractory status and LOT. This facilitates estimation of relative magnitudes of effect modification and allows us to test for effect modification by pooling all evidence of effect modification across trial and treatment comparison.

RHRs were computed for each trial as follows (a formal definition is provided in Supplementary Materials). First, the trial’s strata were sorted to ensure that the order of strata have similar interpretations across trials and are therefore amenable to meta-analysis. For example, LOT strata were sorted from fewest to most LOT, and previous lenalidomide use was nominated as the first (i.e., reference) level of the refractory status factor variable. Then, we computed the ratio between the HR for each stratum and the HR for its preceding stratum (except for the first stratum, which is the reference). Finally, we “inverted” any of these ratios with a point estimate less than one to ensure that point estimates for all RHRs are greater than or equal to one. This inversion step is necessary to prevent ratios less than one from cancelling ratios greater than one in the meta-analyses and thereby obscuring any evidence of effect modification (see below). Standard errors on RHRs were computed as described in Supplementary Materials. We excluded reference strata from meta-analysis because, as references, they are not defined with respect to another stratum.

The RHR scale removes heterogeneity in direction of treatment effect within and between trials and facilitates meta-analysis across all trials such as to make evidence of effect modification statistically detectable; it therefore strongly favors the effect modification hypothesis. A RHR tells us how many times larger a stratified estimate is compared to the estimate for its preceding stratum (or vice versa). If the meta-analytical estimate of mean RHR differs statistically from RHR = 1, then we can reject the null hypothesis of no effect modification.

All meta-analyses were performed on the logarithmic scale. We used random-effects models throughout because there are important differences in the definitions of refractory status and LOT used across the trials which would be expected to manifest as heterogeneity, and which must be accounted for statistically. We present results using forest plots, sub-grouped by publication, to report estimates of mean HRs or mean RHRs, 95% confidence intervals, and *I*^2^ and *p*-values throughout. We used the conventional *p* < 0.05 criterion for statistical significance. Statistical analyses were performed using Stata 16 (StataCorp LLC, College Station, Texas, USA). Data and code are freely available (see the Data and Software Availability section).

### Simulation studies

To help understand the degree to which effect modification may affect NMA results, we performed two simulation studies (plus various sensitivity analyses; see Discussion). The purpose of the simulations was to estimate the percentage of NMA estimates that would be expected to be statistically significantly different under effect modification compared to no effect modification, due to refractory status and LOT. Figure 1 shows a cartoon that illustrates the design of these studies.

**Figure 1:**
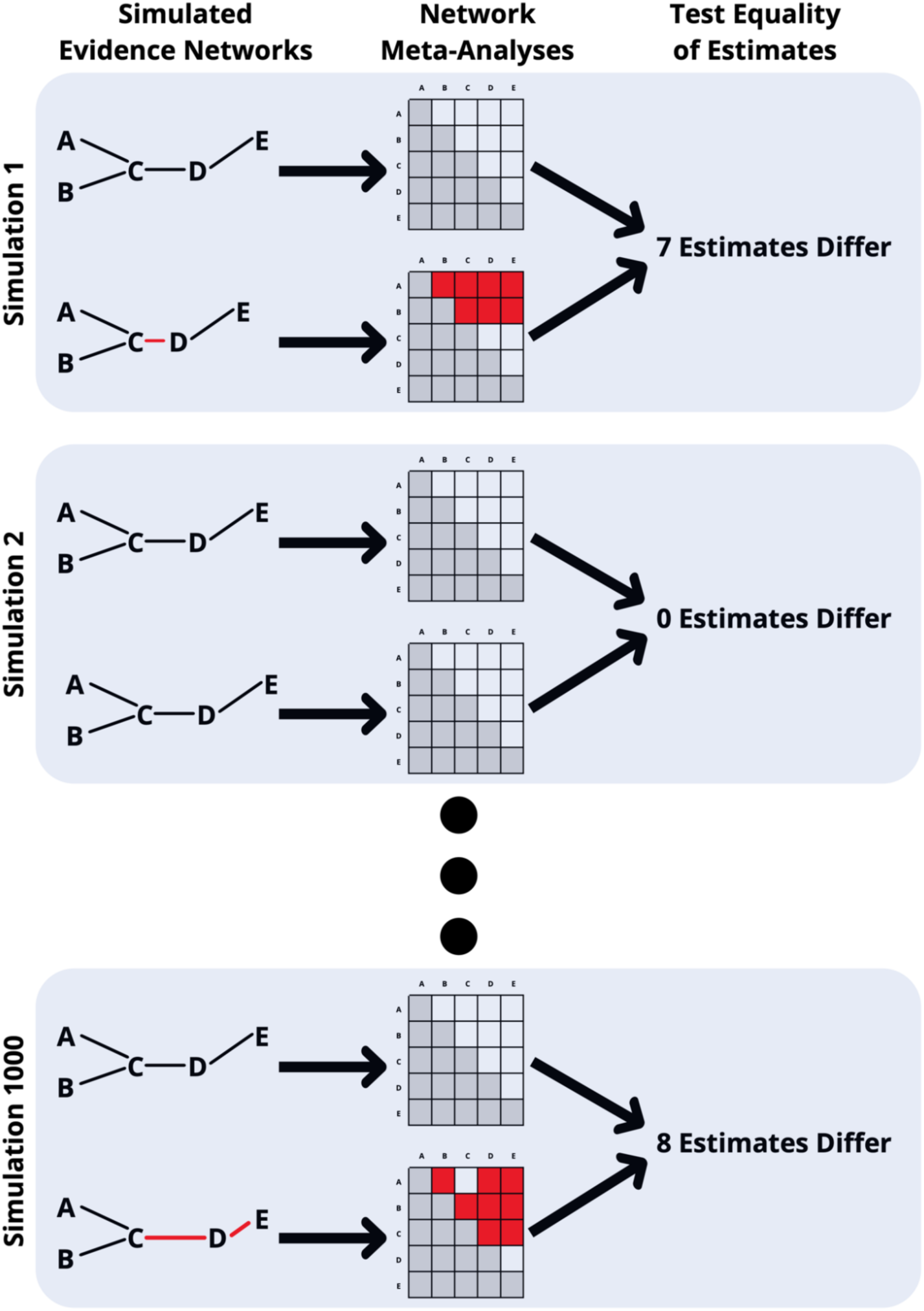
Cartoon of the simulation study. Each panel shows one of the 1000 pairs of simulated networks. Within a pair, the estimates of one network (the topmost in the cartoon) were only subject to simulated heterogeneity, while the other was subject to heterogeneity and effect modification. Each network in the cartoon has 5 treatments (A, B, …, E), but the simulations used 35 treatments. The magnitudes of direct estimates of effect are indicated by the lengths of the links between treatments (heterogeneity and effect modification affect the magnitudes of the estimates, and in extreme cases, their directions). Direct estimates that are particularly modified are shown as red links. NMA results are indicated by the matrices. Diagonal elements are not considered further (shaded) because there is no treatment effect between one treatment and itself. Lower triangles are not considered further (shaded) because they are identical to the upper triangles except for direction (sign). Corresponding estimates within a pair are tested for equality, and those that differ significantly are counted. Effect modification is quite severe in the first and final simulations illustrated by the cartoon, with 7 and 8 estimates differing. No estimates are statistically different in the final simulation. These numbers are merely illustrative. By performing many simulations it is possible to estimate the proportion of NMA estimates that would expected to be affected by the degree of effect modification observed in the literature.

Each simulation used 1000 pairs of synthetic networks of evidence, generated to be similar in distribution to the real network for PFS (the outcome for which RHRs were estimated to be largest; see Results). Networks within a pair were identical except that one network was subjected to simulated effect modification and the other was not, such that any differences in NMA estimates between the two networks could only be attributed to the impact of effect modification. All networks had the same topology as the network for PFS. Simulated effect sizes (log HRs) and their standard errors were drawn from distributions that matched those for the PFS data.

We used estimates of RHR for PFS because they were larger than for OS (i.e., we assumed worst-case scenarios), simulating effect modification by sampling from normal distributions parameterized by mean RHRs and their standard errors to account for uncertainty on the estimates of RHR. We fitted random-effects component-NMA models^25^ to each pair of simulated networks and tested null hypotheses of no differences between corresponding estimates. Testing was performed using two-sided *Z*-tests using the estimated log HRs and their standard errors. Corresponding estimates were deemed to differ if *p* < 0.05. We summarized the results of each simulation as the percentage of estimates expected to be statistically significantly different under effect modification compared to no effect modification. Simulations were performed using R version 3.5.2^26^ with component NMAs performed using the netmeta^27^ package (version 1.3-0). Further details are available in Supplementary Methods.

## RESULTS

Systematic literature searching identified 810 references, of which 40 publications contributed stratified estimates (see Supplementary Materials). Table 1 summarizes the included publications. It shows which trials could have reported stratified estimates (because they included patients that differ with respect to refractory status or LOT) and did so; trials and publications that could have reported stratified estimates but chose not to; and trials that could not report stratified estimates.

**Table 1:** Overview of included publications.

| Publication (Trial) | Comparison<sup>†</sup> | Could report HRs stratified by<sup>*</sup> |  | Does report HRs stratified by<sup>**</sup> |  |  |  |
| --- | --- | --- | --- | --- | --- | --- | --- |
|  |  | Refractory status<sup>††</sup> | Lines of treatment | Refractory status<sup>††</sup> |  | Lines of treatment |  |
|  |  |  |  | OS | PFS | OS | PFS |
| Attal 2019 (ICARIA-MM)<sup>32</sup> | IsPd vs Pd | ✓ | ✓ |  | ✓ |  | ✓ |
| Bahlis 2020 (POLLUX)<sup>33</sup> | DRd vs Rd | ✓ | ✓ |  | ✓ |  | ✓ |
| Dimopoulos 2007 (MM-010)<sup>34</sup> | Rd vs d | ✓ | ✓ |  |  |  |  |
| Dimopoulos 2013 (VANTAGE 088)<sup>35</sup> | VorV vs V | ✓ | ✓ |  | ✓ |  | ✓ |
| Dimopoulos 2016 (ENDEAVOR)<sup>28</sup> | Kd vs Vd | ✓ | ✓ |  | ✓ |  | ✓ |
| Dimopoulos 2018 (ELOQUENT-3)<sup>36</sup> | EPd vs Pd | ✓ | ✓ |  |  |  |  |
| Dimopoulos 2020 (ELOQUENT-2)<sup>37</sup> | ERd vs Rd | ✓ | ✓ |  | ✓ |  | ✓ |
| Dimopoulos 2021 (APOLLO)<sup>38</sup> | DPd vs Pd | ✓ | ✓ |  |  |  |  |
| Dimopoulos 2021<sup>39</sup> | Is vs Isd |  | ✓ |  |  |  |  |
| Garderet 2012 (MMVAR/IFM 2005-04)<sup>40</sup> | VTd vs Td | ✓ |  |  |  |  |  |
| Grosicki 2020 (BOSTON)<sup>41</sup> | SeVd vs Vd | ✓ | ✓ |  | ✓ |  | ✓ |
| Hájek 2017 (FOCUS (PX-171-011))<sup>42</sup> | K vs d | ✓ | ✓ |  |  |  |  |
| Hou 2017 (China Cont. Study)<sup>43</sup> | IRd vs Rd | ✓ | ✓ | ✓ | ✓ | ✓ | ✓ |
| Iida 2018<sup>44</sup> | Vd vs Td |  | ✓ |  |  |  |  |
| Jakubowiak 2016<sup>45</sup> | EVd vs Vd | ✓ | ✓ |  | ✓ |  | ✓ |
| Kropff 2017<sup>46</sup> | Vd vs CyVd | ✓ | ✓ |  |  |  |  |
| Kumar 2020 (BELLINI)<sup>47</sup> | VeVd vs Vd | ✓ | ✓ | ✓ | ✓ | ✓ | ✓ |
| Lonial 2015 (ELOQUENT-2)<sup>48</sup> | Eld vs Ld | ✓ | ✓ |  | ✓ |  |  |
| Lu 2021 (LEPUS)<sup>49</sup> | DVd vs Vd | ✓ | ✓ |  | ✓ |  | ✓ |
| Mateos 2019 (KEYNOTE-183)<sup>50</sup> | PemPd vs Pd | ✓ | ✓ |  |  |  |  |
| Mateos 2020 (CASTOR)<sup>29</sup> | DVd vs Vd | ✓ | ✓ |  | ✓ |  | ✓ |
| Montefusco 2020<sup>51</sup> | CyVd vs CyRd | ✓ |  |  |  |  |  |
| Moreau 2016 (TOURMALINE-MM1)<sup>52</sup> | IRd vs Rd | ✓ | ✓ |  |  |  |  |
| Moreau 2021 (IKEMA)<sup>53</sup> | IsKd vs Kd | ✓ | ✓ |  |  |  |  |
| Orlowski 2007 (DOXIL-MMY-3021)<sup>54</sup> | V vs DoxV | ✓ | ✓ |  | ✓ |  |  |
| Orlowski 2015<sup>55</sup> | SV vs V | ✓ | ✓ |  |  |  |  |
| Orlowski 2016 (DOXIL-MMY-3021)<sup>56</sup> | V vs DoxV | ✓ | ✓ | ✓ |  |  |  |
| Orlowski 2019 (ENDEAVOR)<sup>57</sup> | Kd vs Vd | ✓ | ✓ | ✓ |  | ✓ |  |
| Palumbo 2016 (CASTOR)<sup>58</sup> | DVd vs Vd | ✓ | ✓ |  | ✓ |  | ✓ |
| Raje 2017<sup>59</sup> | TabVd vs Vd | ✓ | ✓ |  |  |  |  |
| Richardson 2007 (APEX)<sup>60</sup> | V vs d | ✓ | ✓ |  |  |  |  |
| Richardson 2014 (MM-02)<sup>61</sup> | Pd vs P | ✓ | ✓ |  |  |  |  |
| Richardson 2019 (OPTIMISMM)<sup>62</sup> | PVd vs Vd | ✓ | ✓ |  | ✓ |  | ✓ |
| Richardson 2021 (TOURMALINE-MM1)<sup>63</sup> | IRd vs Rd | ✓ | ✓ | ✓ |  | ✓ |  |
| San-Miguel 2013 (MM-03)<sup>64</sup> | Pd vs d | ✓ | ✓ |  |  | ✓ | ✓ |
| San-Miguel 2014 (PANORAMA-1)<sup>65</sup> | FVd vs Vd | ✓ | ✓ |  |  |  | ✓ |
| San-Miguel 2016 (PANORAMA-1)<sup>66</sup> | FVd vs Vd | ✓ | ✓ | ✓ |  | ✓ |  |
| Siegel 2018 (ASPIRE)<sup>67</sup> | KRd vs Rd | ✓ | ✓ |  |  | ✓ |  |
| Stewart 2015 (ASPIRE)<sup>68</sup> | KRd vs Rd | ✓ | ✓ |  | ✓ |  |  |
| Usmani 2022 (CANDOR)<sup>69</sup> | DKd vs Kd | ✓ | ✓ |  | ✓ |  | ✓ |
| Weber 2007 (MM-09)<sup>70</sup> | Rd vs d | ✓ | ✓ |  |  |  |  |
| White 2013 (AMBER)<sup>71</sup> | BevV vs V | ✓ | ✓ |  |  |  |  |
| **Publications (%)** | **42** | **40 (95%)** | **40 (95%)** | **6 (14%)** | **17 (40%)** | **7 (17%)** | **18 (43%)** |

\*Trials that include only refractory patients, or patients who received a specific number of previous lines of treatment, cannot report stratified estimates. Determinations were made using tables of baseline characteristics and inclusion criteria.
\*\*Publications that report stratified estimates graphically rather than numerically are classified as not reporting stratified estimates. Publications that report a stratified estimate for only one stratum (e.g., refractory but not also non-refractory) are classified as not reporting stratified estimates as they provide incomplete information. It would not be practical or possible for some trials to report stratified estimates due to very small sample sizes or no or very few events.
†Treatment names are listed in full in Supplementary Materials.
††A broad definition of refractory status or previous treatment use was used (see Methods).

### Effect modification of HR for progression-free survival

Almost all trials could have reported stratified estimates, but only 17 (40%) publications, representing 8364 patients, did report estimates stratified by refractory status (Table 1). Similarly, 18 (43%) publications, representing 7,503 patients, did report estimates stratified by LOT (Table 1). Within-trial evidence for effect modification of HR for PFS by refractory status and LOT is weak (Figure 2 and Figure 3). Only one test for equality of stratified HRs was statistically significant with respect to refractory status (*p* < 0.01 for the comparison Kd vs Vd^28^) and another with respect to LOT (*p* = 0.01 for the comparison DVd vs Vd^29^).

**Figure 2:**
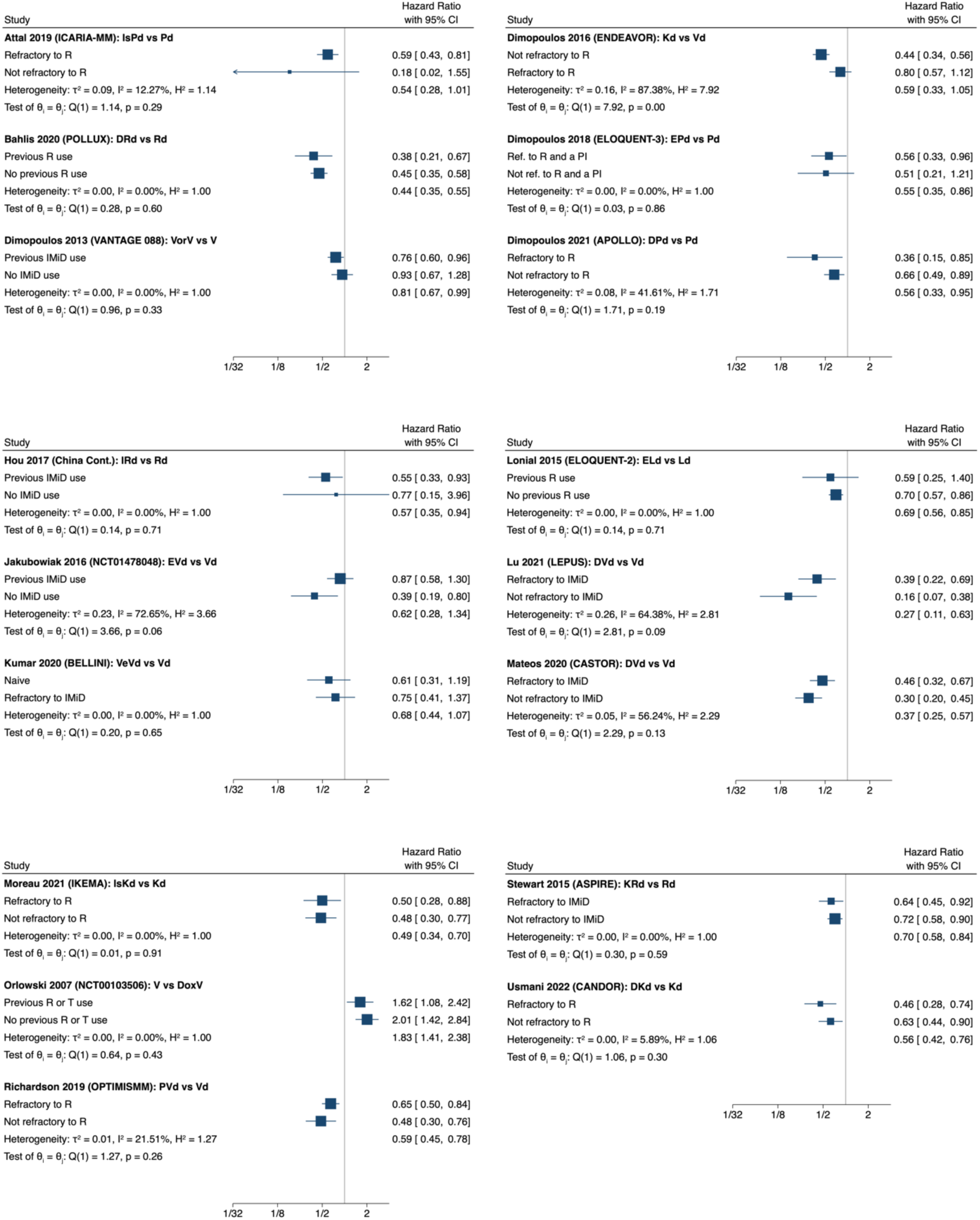
Hazard ratios for PFS stratified by refractory status. Statistically significant stratified estimates of HR indicate likely treatment effect in specific patient subgroups. Effect modification would be demonstrated by unequal stratified HRs within trial. Only one of the 17 within-trial tests for equality of stratified HRs give statistically significant results at the 95% significance level. Note the lack of a consistent pattern in the estimates across trial that would lend face-validity to the effect modification hypothesis.

**Figure 3:**
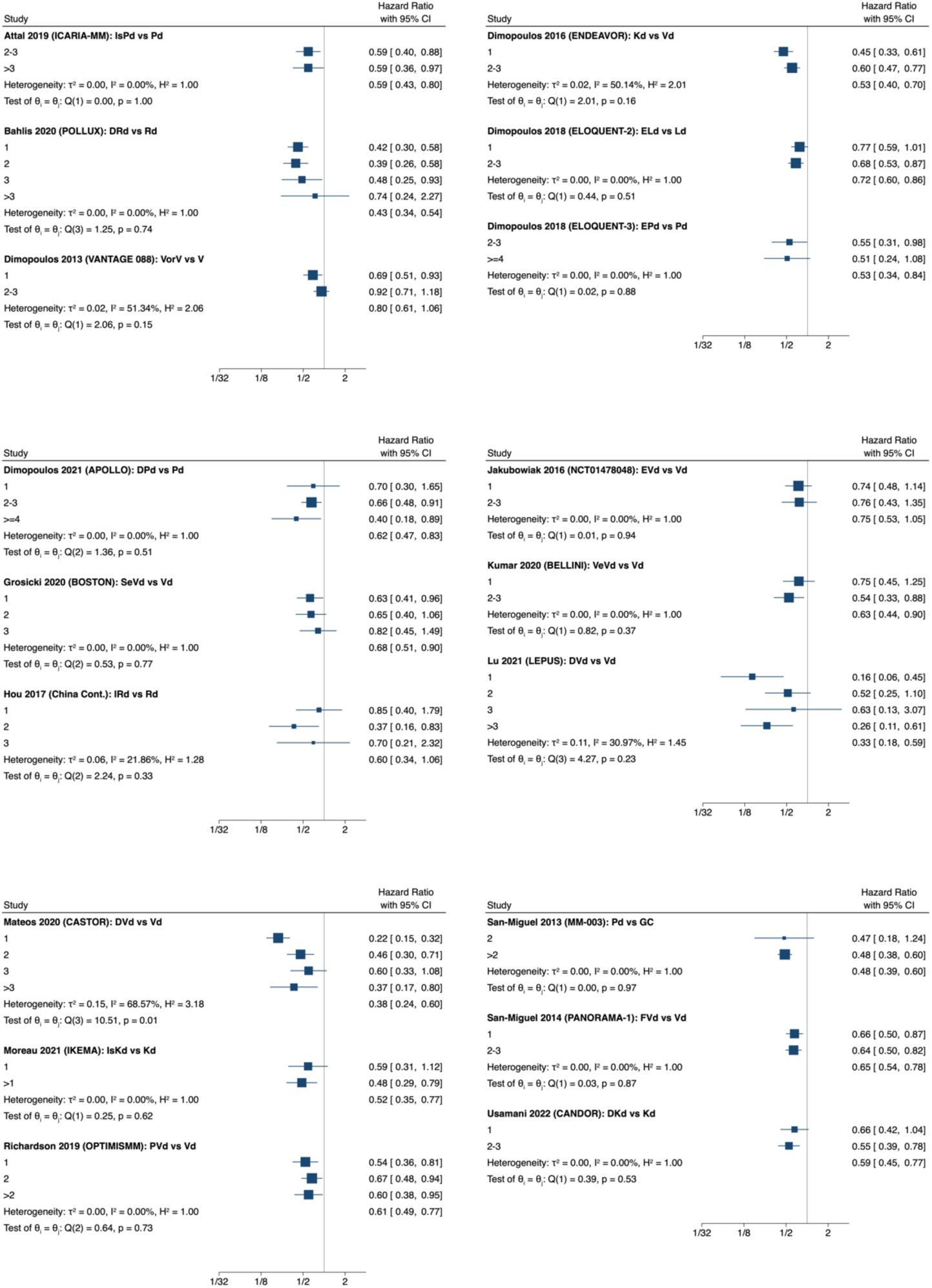
Hazard ratios for PFS stratified by number of lines of. Statistically significant stratified estimates of HR indicate likely treatment effect in specific patient subgroups. Effect modification would be demonstrated by unequal stratified HRs within trial. Only one of the 18 within-trial tests for equality of stratified HRs give statistically significant results at the 95% significance level. Note the lack of a consistent pattern in the estimates across trial that would lend face-validity to the effect modification hypothesis.

Mean RHR was estimated to be 1.31 (95% CI 1.16 to 1.47; *p* < 0.005; *I*^2^ = 0%) for refractory status and 1.19 (95% CI 1.09 to 1.29; *p* < 0.01; *I*^2^ = 0%) for LOT (Figure 4). No statistical heterogeneity in RHR was observed.

**Figure 4:**
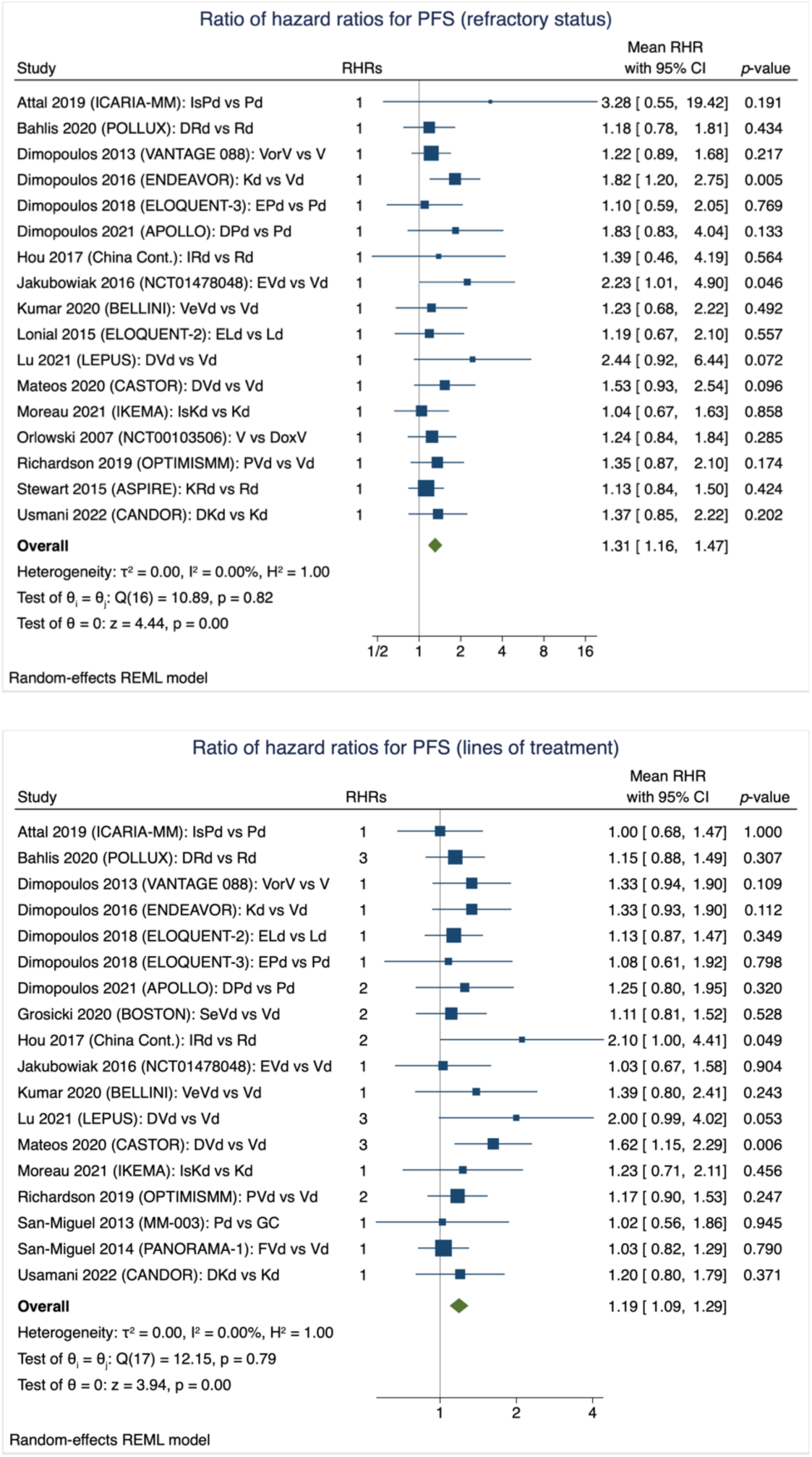
Estimates of ratios of hazard ratios for. The panels show estimates of ratios of hazard ratios (RHRs) constructed under conditions that favor the effect modification hypothesis. The top panel shows RHRs for refractory status and the bottom panel shows RHRs for number of lines of treatment. RHR = 1 corresponds to no effect modification.

### Effect modification of HR for overall survival

Almost all trials could have reported stratified estimates for OS. Only 6 publications (14%), representing 3,471 patients, did report estimates stratified by refractory status (Table 1). Similarly, only 7 (17%) publications, representing 4,063 patients, did report estimates stratified by LOT (Table 1). Within-trial evidence for effect modification of HR for OS by refractory status and LOT is very weak, with no tests for equality of stratified HRs demonstrating statistical significance (Supplementary Materials).

Mean RHR was estimated to be 1.16 (95% CI 1.01 to 1.32; *p* = 0.03; *I*^2^ = 0%) for refractory status and 1.09 (95% CI 0.98 to 1.20; *p* = 0.12; *I*^2^ = 0%) for LOT (Supplementary Materials). No statistical heterogeneity in RHR was observed, suggesting that effect modification may be relatively consistent across trial and comparison, and that our broad definitions of refractory status and LOT did not introduce undue heterogeneity.

### Simulation study

Table 2 summarizes the results for the simulations. We would expect no more than 0.4% of NMA estimates for PFS to differ if trial estimates are, versus are not, subject to effect modification by refractory status. I.e., among the 595 possible comparisons of the 35 treatments in the included trials on PFS, we would expect no more than about 2–3 comparisons to differ statistically significantly due to effect modification. We would expect no more than 5% of NMA estimates for PFS to differ if trial estimates are, versus are not, subject to effect modification by LOT. I.e., among the 595 possible comparisons, no more than about 30 comparisons would be expected to differ statistically significantly due to effect modification.

**Table 2:** Simulation study results: Percentages of NMA estimates for progression free survival expected to differ in the presence of effect modification and heterogeneity.

| Potential effect modifier | NMA estimates expected to differ (95% CI) |
| --- | --- |
| Refractory status* | 0.35% (0.33%–0.36%) |
| Lines of Treatment† | 4.48% (4.42%–4.53%) |

\*A broad definition of refractory status or previous treatment use was used (see Methods). †Results for lines of treatment assume four categories, that effect modification consistently increases or decreases with LOT, and that effect modification compounds over categories (see Methods).

## DISCUSSION

We have systematically reviewed and meta-analyzed the evidence for effect modification of HR for OS and PFS stratified by refractory status and LOT. Within-trial evidence for effect modification of HR for OS and PFS by refractory status and LOT is weak, with only 2 of 49 tests of heterogeneity demonstrating statistical significance (i.e., almost exactly the number of type I errors expected at the 95% significance level under the null hypothesis of no effect modification). Of the meta-analyses of cross-trial effect modification, the largest (i.e., worst-case) mean RHR estimated was 1.31 (95% CI 1.16 to 1.47) for HR for PFS with respect to refractory status.

We then used worst-case estimates of RHR in simulations to estimate percentages of NMA estimates that may be affected by effect modification. For refractory status, the simulations suggest that even if effect modification is as large as the worst-case estimate, substantially fewer than 1% of NMA estimates are likely to be statistically different than they would be if effect modification does not occur. For LOT, the simulations suggest that fewer than 5% of NMA estimates are likely to be statistically different in the presence of effect modification and heterogeneity. This is substantially higher than for refractory status, but putting this result in perspective, 5% is the same as our typical tolerance for type I errors given by the conventional significance level of 95%.

Absence of evidence is not evidence of absence, and we may simply not have sufficient data to detect the impact of effect modification. Still, if effect modification does occur, we would expect to see consistent patterns which support effect modification, which we do not. In some cases, estimates increase with refractory status or LOT, in others it is opposite, but in most cases the estimates are practically the same.

Cope et al. qualitatively assessed 12 NMAs or unanchored indirect comparisons and, based on expert opinions on variables that may be effect modifiers, concluded that the NMA estimates may have been compromised by differences in distributions of effect modifiers.^13^ We are aware of one other attempt at quantifying effect modification through meta-analysis of results from subgroups, e.g., patients with one previous LOT vs. 2 or more previous LOT.^6^ In this NMA, which was limited to comparing different immunomodulatory-containing regimens for RRMM, Dimopoulos et al. reported that their subgroup analyses yielded results consistent with their main findings (i.e., no apparent effect modification).

### Strengths and limitations

To our knowledge, this is the first systematic review, meta-analysis, and simulation study of effect modification in RRMM. However, it was not prespecified. While we were able to obtain stratified estimates from up to 18 (43%) of the included publications, most publications did not report stratified estimates. Because we did not perform a separate literature search, it is possible that we do not have all the available data on effect modification. However, it would be unreasonable to expect the smaller trials to report stratified estimates, as they would likely be so imprecise as to be uninformative. Further, stratified estimates were published for about half as many analyses of OS compared to PFS, despite there being about the same number of publications providing estimates of HR for the two outcomes. Because stratified estimates are not reported in the main trial reports for so many comparisons, it is possible that effect modification is larger than we estimate, particularly for OS.

While we systematically reviewed evidence of effect modification with respect to refractory status and number of lines of treatment, we did not systematically review other variables that also may be effect modifiers. However, we did look at all stratified estimates when extracting data and did not notice any variables that appeared to consistently demonstrate convincing evidence of effect modification.

Because there was heterogeneity in trial reporting, we were not able to use definitions of refractory status and number of lines of treatment that measured exactly what we were interested in, because doing so would have resulted in almost no synthesizable evidence. We therefore used pragmatic and inclusive definitions, particularly for refractory status (see Methods). We would have expected this to introduce heterogeneity, but this was not statistically observable in the meta-analyses of RHR (*I*^2^ = 0% in all analyses).

Because the within-trial evidence of effect modification is so weak, but there are nevertheless concerns in the RRMM research community about effect modification and NMA, we constructed RHR and designed the simulations to strongly favor the effect modification hypothesis. This likely resulted in exaggerated conclusions about whether effect modification occurs and the extent to which it is problematic.

Quantities such as ratios of hazard ratios, as used in meta-research,^30^ are likely challenging to interpret, and we suspect that few will have an intuitive understanding of what constitutes a “large” or “important” RHR with respect to effect modification in RRMM. A major strength of this work is that having estimated RHRs, we then used simulations to investigate how many NMA estimates would be expected to be statistically significantly different under the estimated degree of effect modification. We hope this helps readers understand the likely impact of any effect modification on NMA estimates. However, we remind readers that we used random-effects NMAs,^31^ which are designed to account for heterogeneity in trial estimates. Our results do not necessarily translate to fixed-effects NMAs, as used in some systematic reviews on treatments for RRMM.^4,2^ It is important to note that fixed- and random-effects NMAs make fundamentally different assumptions about transitivity. Fixed-effects NMAs assume that “trial-level” treatment effects can be added and subtracted to make indirect estimates. Random-effects NMAs assume that *average* treatment effects can be added and subtracted. Use of average treatment effects explicitly accounts for differences between trials, including different distributions of effect modifiers.

Finally, we also performed sensitivity analyses to investigate the implications of the assumptions we made in the simulations. For example, because RHR discards direction of modification, we assumed that direction of modification is consistent within treatment comparison but may vary between comparisons. This may not be true, so we performed a sensitivity analysis in which direction is assumed consistent within and between comparisons. The result of this analysis suggests that about half as many estimates would differ statistically compared to the main analysis (i.e., that the main result reflects the worst-case).

### Implications for research

Understandably, the transitivity assumption underpinning NMA is often highly simplified in articles aimed at non-statisticians. For example, articles tend to use arguments about “similarity” of patients.^8^ Given this oversimplification, it is unsurprising there are concerns about using NMA in RRMM and other areas. The transitivity assumption does not concern patient similarity, nor whether treatment effect *estimates* can be added or subtracted, it concerns whether target parameters can be linearly combined. Patient similarity is a good place to start thinking about NMAs, but a terrible place to stop. Modern statistical methods should be communicated more carefully and received more studiously.

Understanding effect modification is important for making decisions based on individual trials, and for assessing the assumptions and validity of NMAs. We therefore suggest that RRMM trialists develop and adopt standardized definitions of potential effect modifiers that, where possible, should be used to report stratified analyses in future trials. In addition to improving transparency and improving consistency of reporting of effect estimates for patient subgroups, standardization would facilitate more specific meta-analytical study of effect modification by reducing methodological heterogeneity. Further, we suggest that stratified analyses be reported for all patient-important outcomes, particularly OS, which has been dramatically underreported compared to PFS.

The strength of concerns that effect modification, as it may occur in RRMM, may invalidate NMAs appears to be inconsistent with the available evidence. This suggests that NMA can probably be relied upon to estimate direct and indirect treatment effects, subject to some important caveats. First, evidence on effect modification is limited to at most ∼40% of comparisons, so it is possible that modification is more severe in the remaining ∼60% of comparisons. That said, it would be concerning if large modification occurs but has been systematically unreported in the majority of phase 2 and 3 trials. Second, more evidence on modification is available for PFS than OS, so it is possible that HR for OS is subject to greater modification than the available evidence suggests. This may be because the PFS endpoint is typically reached earlier than that for OS. However, again, it would be concerning if large modification was not being reported for what is arguably the most important outcome of cancer trials. Third, we are not suggesting that a particular NMA estimate can be applied to patients who are refractory to one or both treatments involved in a given comparison: such estimates would be subject to an obvious, if somewhat absurd, form of effect modification. A method for ranking treatments for patients who are refractory to specific treatments or components is presented in the Supplementary Appendix. Fourth, and crucially, our simulations estimated random-effects (cf. fixed-effects). Random-effects models account for heterogeneity in effect estimates due not just to sampling error, but also other factors, including effect modification. Our findings are unlikely to generalize to fixed-effects NMAs. Finally, NMA should be able to be used to make indirect estimates if effect modification is negligible for all direct comparisons and there is no good reason to believe that non-negligible modification would occur for treatment comparisons that have not been made directly. However, it would be preferable to have direct evidence.

### Summary and conclusions

There is very weak within-trial evidence for effect modification with respect to refractory status and number of previous lines of treatment. It is plausible that effect modification does not occur with respect to these variables or is so small as to be statistically undetectable, even in phase 3 trials. If this is true, then differences in the distributions of these variables across trials are unlikely to be a problem in NMAs. We were only able to detect effect modification by estimating ratios of hazard ratios (RHRs) across trials under assumptions that strongly favor the modification hypothesis. These assumptions may not hold, so our estimates of the magnitude of effect modification may be exaggerated, as may our estimates of the percentages of NMA estimates that would be expected to be affected.

Adequately performed random-effects NMAs can probably be relied upon to provide direct and indirect estimates of mean HRs for OS and PFS, subject to the caveats discussed above.

## Data Availability

Data and software are publicly available at: https://github.com/multinormal/fhi.rrmm-em.2022
The specific version used to generate the results presented herein is archived at Zenodo: https://doi.org/10.5281/zenodo.6545584

https://doi.org/10.5281/zenodo.6545584

## AUTHOR CONTRIBUTIONS

Study conception and design: CJR

Acquisition, analysis, or interpretation of data: CJR, IKO, LG, GEN, AF

Drafting the manuscript: CJR, IKO, GEN, AF

Critical revision of the manuscript for important intellectual content: IKO, LG, GEN, AF

Statistical analysis: CJR

Supervision: AF

All authors approved the published version and agreed to be accountable for all aspects of the work.

## DATA AND SOFTWARE AVAILABILITY

Data and software are publicly available at: https://github.com/multinormal/fhi.rrmm-em.2022 The specific version used to generate the results presented herein is archived at Zenodo: https://doi.org/10.5281/zenodo.6545584

## DISCLOSURE OF CONFLICTS OF INTEREST AND FUNDING

None of the authors have any competing interests with respect to this work.

The authors conducted this research under the employ of the Norwegian Institute of Public Health (Folkehelseinstituttet). The work was funded via Norway’s National System for Managed Introduction of New Health Technologies within the Specialist Health Service (Nye Metoder). The funder had no role in the conduct of this work.

## SUPPLEMENTARY MATERIAL

The following supplementary material accompanies this manuscript:

1. Supplementary Materials.docx

## Notes

### Competing Interest Statement

The authors have declared no competing interest.

### Author Declarations

The study used only previously published data.

### Summary of Updates

This version corrects a typo in eq. 6 of supplementary materials.

