## Supplementary Materials for "Effect modification in network meta-analyses of treatments for relapsing refractory multiple myeloma (RRMM): systematic review, meta-analysis, and simulation"

**Working Paper** — This document has not yet been submitted for peer review and may be revised.

Search strategy from February-August 2020 ... 14

Search strategy from August 2020 – March 2021 ... 17

Search strategy for ongoing studies ... 20

**Missing data ... 21**

**Further details on the simulation study ... 21**

**Ratio of hazard ratios and its sampling variance ... 23**

***Appendix: Conditional P-scores for treatment ranking when some treatments are not competitors ... 24***

**Introduction ... 24**

**Theory ... 25**

**References ... 26**

### Supplementary Tables

#### *Supplementary Table 1: Included treatments and their abbreviations*

Note: Not all treatments are included in meta-analyses for OS and PFS.

| Treatment | Abbreviation |
|---|---|
| Observation | Observation |
| Bevacizumab and bortezomib | BevV |
| Bortezomib, cyclophosphamide and dexamethasone | CyVd |
| Bortezomib and dexamethasone | Vd |
| Bortezomib | V |
| Bortezomib and pegylated liposomal doxorubicin | DoxV |
| Bortezomib, thalidomide and dexamethasone | VTd |
| Bortezomib and vorinostat | VorV |
| Carfilzomib and dexamethasone | Kd |
| Carfilzomib, lenalidomide and dexamethasone | KRd |
| Carfilzomib | K |
| Glucocorticoid (unspecified) | d |
| Daratumumab and methylprednisone | Dd |
| Daratumumab, bortezomib and dexamethasone | DVd |
| Daratumumab, carfilzomib and dexamethasone | DKd |
| Daratumumab, lenalidomide and dexamethasone | DRd |
| Daratumumab, pomalidomide and dexamethasone | DPd |
| Daratumumab | D |
| Dexamethasone | d |
| Elotuzumab, bortezomib and dexamethasone | EVd |
| Elotuzumab, lenalidomide and dexamethasone | ERd |
| Elotuzumab, pomalidomide and dexamethasone | EPd |
| Isatuximab | Is |
| Isatuximab and dexamethasone | Isd |
| Isatuximab, carfilzomib, and dexamethasone | IsKd |
| Isatuximab, pomalidomide and dexamethasone | IsPd |
| Ixazomib, lenalidomide and dexamethasone | IRd |
| Lenalidomide, cyclophosphamide and dexamethasone | CyRd |
| Lenalidomide and dexamethasone | Rd |
| Panobinostat, bortezomib and dexamethasone | FVd |
| Pembrolizumab, pomalidomide and dexamethasone | PemPd |
| Pomalidomide, bortezomib and dexamethasone | PVd |
| Pomalidomide, cyclophosphamide and dexamethasone | CyPd |
| Pomalidomide and dexamethasone | Pd |
| Pomalidomide | P |

| | |
|---|---|
| Siltuximab and bortezomib | SV |
| Selinexor, bortezomib and dexamethasone | SeVd |
| Tabalumab, bortezomib and dexamethasone | TabVd |
| Thalidomide and dexamethasone | Td |
| Venetoclax, bortezomib and dexamethasone | VenVd |

### *Supplementary Table 2: PRISMA 2020 Checklist*

| Section and Topic | Item # | Checklist item | Location where item is reported |
|---|---|---|---|
| **TITLE** | | | |
| Title | 1 | Identify the report as a systematic review. | Title |
| **ABSTRACT** | | | |
| Abstract | 2 | See the PRISMA 2020 for Abstracts checklist. | SEE NOTE 1 |
| **INTRODUCTION** | | | |
| Rationale | 3 | Describe the rationale for the review in the context of existing knowledge. | Introduction |
| Objectives | 4 | Provide an explicit statement of the objective(s) or question(s) the review addresses. | Introduction |
| **METHODS** | | | |
| Eligibility criteria | 5 | Specify the inclusion and exclusion criteria for the review and how studies were grouped for the syntheses. | Methods |
| Information sources | 6 | Specify all databases, registers, websites, organisations, reference lists and other sources searched or consulted to identify studies. Specify the date when each source was last searched or consulted. | This document and SEE NOTE 2 |
| Search strategy | 7 | Present the full search strategies for all databases, registers and websites, including any filters and limits used. | This document |
| Selection process | 8 | Specify the methods used to decide whether a study met the inclusion criteria of the review, including how many reviewers screened each record and each report retrieved, whether they worked independently, and if applicable, details of automation tools used in the process. | SEE NOTE 2 |
| Data collection process | 9 | Specify the methods used to collect data from reports, including how many reviewers collected data from each report, whether they worked independently, any processes for obtaining or confirming data from study investigators, and if applicable, details of automation tools used in the process. | Methods |
| Data items | 10a | List and define all outcomes for which data were sought. Specify whether all results that were compatible with each outcome domain in each study were sought (e.g. for all measures, time points, analyses), and if not, the methods used to decide which results to collect. | Methods |
| | 10b | List and define all other variables for which data were sought (e.g. participant and intervention characteristics, funding sources). Describe any assumptions made about any missing or unclear information. | Methods |
| Study risk of bias assessment | 11 | Specify the methods used to assess risk of bias in the included studies, including details of the tool(s) used, how many reviewers assessed each study and whether they worked | SEE NOTE 2 |

| Section and Topic | Item # | Checklist item | Location where item is reported |
|---|---|---|---|
| | | independently, and if applicable, details of automation tools used in the process. | |
| Effect measures | 12 | Specify for each outcome the effect measure(s) (e.g. risk ratio, mean difference) used in the synthesis or presentation of results. | Methods |
| Synthesis methods | 13a | Describe the processes used to decide which studies were eligible for each synthesis (e.g. tabulating the study intervention characteristics and comparing against the planned groups for each synthesis (item #5)). | Methods |
| | 13b | Describe any methods required to prepare the data for presentation or synthesis, such as handling of missing summary statistics, or data conversions. | Methods and Supplementary Methods |
| | 13c | Describe any methods used to tabulate or visually display results of individual studies and syntheses. | Methods |
| | 13d | Describe any methods used to synthesize results and provide a rationale for the choice(s). If meta-analysis was performed, describe the model(s), method(s) to identify the presence and extent of statistical heterogeneity, and software package(s) used. | Methods |
| | 13e | Describe any methods used to explore possible causes of heterogeneity among study results (e.g. subgroup analysis, meta-regression). | NA |
| | 13f | Describe any sensitivity analyses conducted to assess robustness of the synthesized results. | Methods |
| Reporting bias assessment | 14 | Describe any methods used to assess risk of bias due to missing results in a synthesis (arising from reporting biases). | NA |
| Certainty assessment | 15 | Describe any methods used to assess certainty (or confidence) in the body of evidence for an outcome. | NA |
| **RESULTS** | | | |
| Study selection | 16a | Describe the results of the search and selection process, from the number of records identified in the search to the number of studies included in the review, ideally using a flow diagram. | Results |
| | 16b | Cite studies that might appear to meet the inclusion criteria, but which were excluded, and explain why they were excluded. | SEE NOTE 3 |
| Study characteristics | 17 | Cite each included study and present its characteristics. | SEE NOTE 4 |
| Risk of bias in studies | 18 | Present assessments of risk of bias for each included study. | SEE NOTE 5 |
| Results of individual studies | 19 | For all outcomes, present, for each study: (a) summary statistics for each group (where appropriate) and (b) an effect estimate and its precision (e.g. confidence/credible interval), ideally using structured tables or plots. | Results |
| Results of syntheses | 20a | For each synthesis, briefly summarise the characteristics and risk of bias among contributing studies. | SEE NOTE 5 |
| | 20b | Present results of all statistical syntheses conducted. If meta-analysis was done, present for each the summary estimate and its precision (e.g. confidence/credible interval) and measures of statistical heterogeneity. If comparing groups, describe the direction of the effect. | Results and Supplementary Materials |
| | 20c | Present results of all investigations of possible causes of heterogeneity among study results. | NA |

| Section and Topic | Item # | Checklist item | Location where item is reported |
|---|---|---|---|
| | 20d | Present results of all sensitivity analyses conducted to assess the robustness of the synthesized results. | Results |
| Reporting biases | 21 | Present assessments of risk of bias due to missing results (arising from reporting biases) for each synthesis assessed. | Results and Discussion |
| Certainty of evidence | 22 | Present assessments of certainty (or confidence) in the body of evidence for each outcome assessed. | NA |
| **DISCUSSION** | | | |
| Discussion | 23a | Provide a general interpretation of the results in the context of other evidence. | Discussion |
| | 23b | Discuss any limitations of the evidence included in the review. | Discussion |
| | 23c | Discuss any limitations of the review processes used. | Discussion |
| | 23d | Discuss implications of the results for practice, policy, and future research. | Discussion |
| **OTHER INFORMATION** | | | |
| Registration and protocol | 24a | Provide registration information for the review, including register name and registration number, or state that the review was not registered. | SEE NOTE 6 |
| | 24b | Indicate where the review protocol can be accessed, or state that a protocol was not prepared. | Methods |
| | 24c | Describe and explain any amendments to information provided at registration or in the protocol. | NA |
| Support | 25 | Describe sources of financial or non-financial support for the review, and the role of the funders or sponsors in the review. | Disclosure of conflicts of interest and funding |
| Competing interests | 26 | Declare any competing interests of review authors. | Disclosure of conflicts of interest and funding |
| Availability of data, code and other materials | 27 | Report which of the following are publicly available and where they can be found: template data collection forms; data extracted from included studies; data used for all analyses; analytic code; any other materials used in the review. | Data and software |

*From:* Page MJ, McKenzie JE, Bossuyt PM, Boutron I, Hoffmann TC, Mulrow CD, et al. The PRISMA 2020 statement: an updated guideline for reporting systematic reviews. BMJ 2021;372:n71. doi: 10.1136/bmj.n71

Notes:

1. See Supplementary Table 3: PRISMA 2020 Abstracts Checklist below.
2. This information is provided in the HTA's protocol, referenced in the manuscript; full details will be provided in the published HTA.

3. Information on studies that might appear to meet the inclusion criteria for the HTA but were excluded will be presented in the published HTA. Table 1 shows which RCTs could be included in the meta-analyses for the present work, and why.
4. Full study characteristics are not relevant to this review but will be reported in full in the published HTA.
5. Risk of bias assessments will be reported in full in the published HTA.
6. The present work was not prespecified or registered. A protocol for the HTA has been published and is referenced.

### *Supplementary Table 3: PRISMA 2020 Abstracts Checklist*

| Section and Topic | Item # | Checklist item | Reported (Yes/No) |
|---|---|---|---|
| **TITLE** | | | |
| Title | 1 | Identify the report as a systematic review. | YES |
| **BACKGROUND** | | | |
| Objectives | 2 | Provide an explicit statement of the main objective(s) or question(s) the review addresses. | YES |
| **METHODS** | | | |
| Eligibility criteria | 3 | Specify the inclusion and exclusion criteria for the review. | SEE NOTE 1 |
| Information sources | 4 | Specify the information sources (e.g. databases, registers) used to identify studies and the date when each was last searched. | SEE NOTE 2 |
| Risk of bias | 5 | Specify the methods used to assess risk of bias in the included studies. | SEE NOTE 2 |
| Synthesis of results | 6 | Specify the methods used to present and synthesise results. | YES |
| **RESULTS** | | | |
| Included studies | 7 | Give the total number of included studies and participants and summarise relevant characteristics of studies. | SEE NOTE 3 |
| Synthesis of results | 8 | Present results for main outcomes, preferably indicating the number of included studies and participants for each. If meta-analysis was done, report the summary estimate and confidence/credible interval. If comparing groups, indicate the direction of the effect (i.e. which group is favoured). | YES |
| **DISCUSSION** | | | |
| Limitations of evidence | 9 | Provide a brief summary of the limitations of the evidence included in the review (e.g. study risk of bias, inconsistency and imprecision). | SEE NOTE 2 |
| Interpretation | 10 | Provide a general interpretation of the results and important implications. | YES |
| **OTHER** | | | |
| Funding | 11 | Specify the primary source of funding for the review. | SEE NOTE 4 |
| Registration | 12 | Provide the register name and registration number. | SEE NOTE 5 |

*From:* Page MJ, McKenzie JE, Bossuyt PM, Boutron I, Hoffmann TC, Mulrow CD, et al. The PRISMA 2020 statement: an updated guideline for reporting systematic reviews. BMJ 2021;372:n71. doi: 10.1136/bmj.n71

Notes:

1. Inclusion criteria are implied by the statement about phase 1 and 2 studies.
2. This information is not reported in the abstract due to space limitations. It is reported in the manuscript.
3. The abstract reports numbers of studies, but numbers of randomized patients are not reported in the abstract due to the word limit. These data are reported in the manuscript.

4. Funding details are reported in the manuscript.

5. The review was not prespecified or registered. This is reported in the manuscript.

### Supplementary Figures

#### *Supplementary Figure 1: Flow diagram showing results of screening and data extraction*

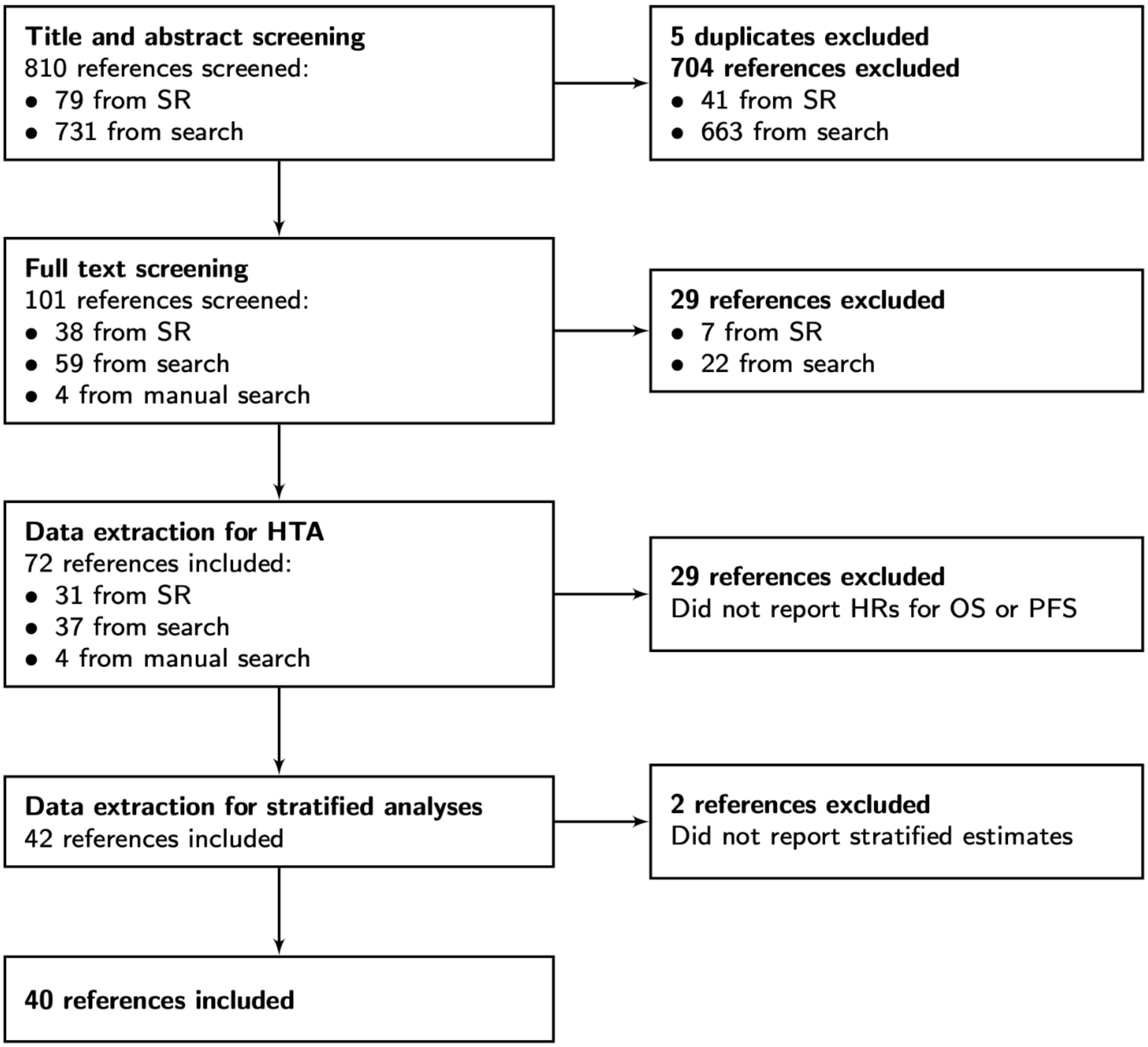

***Supplementary Figure 2: Hazard ratios for OS stratified by refractory status***

Statistically significant stratified estimates of HR indicate likely treatment effect in specific patient subgroups. Effect modification would be demonstrated by unequal stratified HRs within trial. None of the 6 within-trial tests for equality of stratified HRs give statistically significant results at the 95% significance level.

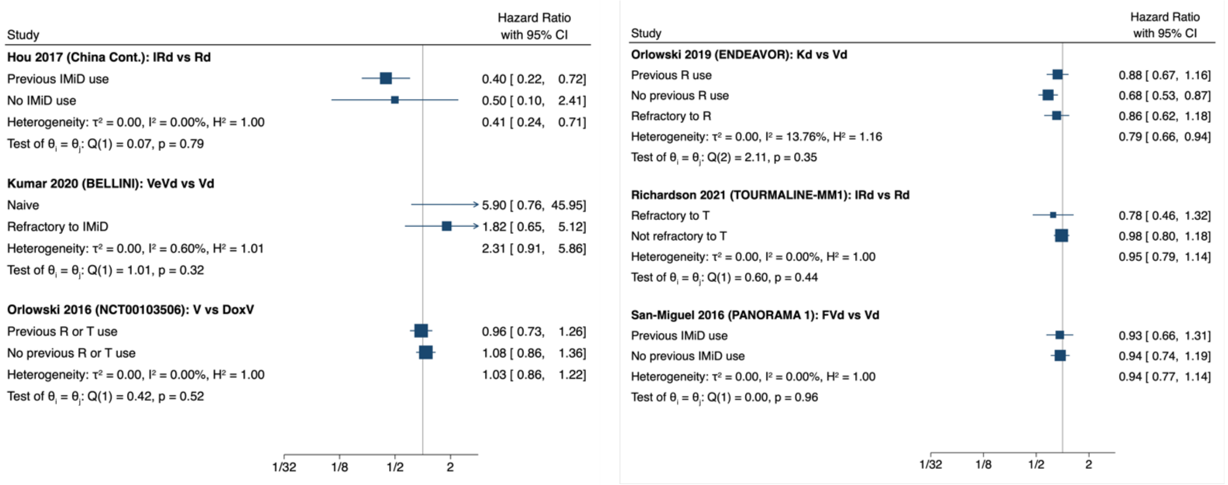

### *Supplementary Figure 3: Hazard ratios for OS stratified by number of lines of treatment*

Statistically significant stratified estimates of HR indicate likely treatment effect in specific patient subgroups. Effect modification would be demonstrated by unequal stratified HRs within trial. None of the 8 within-trial tests for equality of stratified HRs give statistically significant results at the 95% significance level.

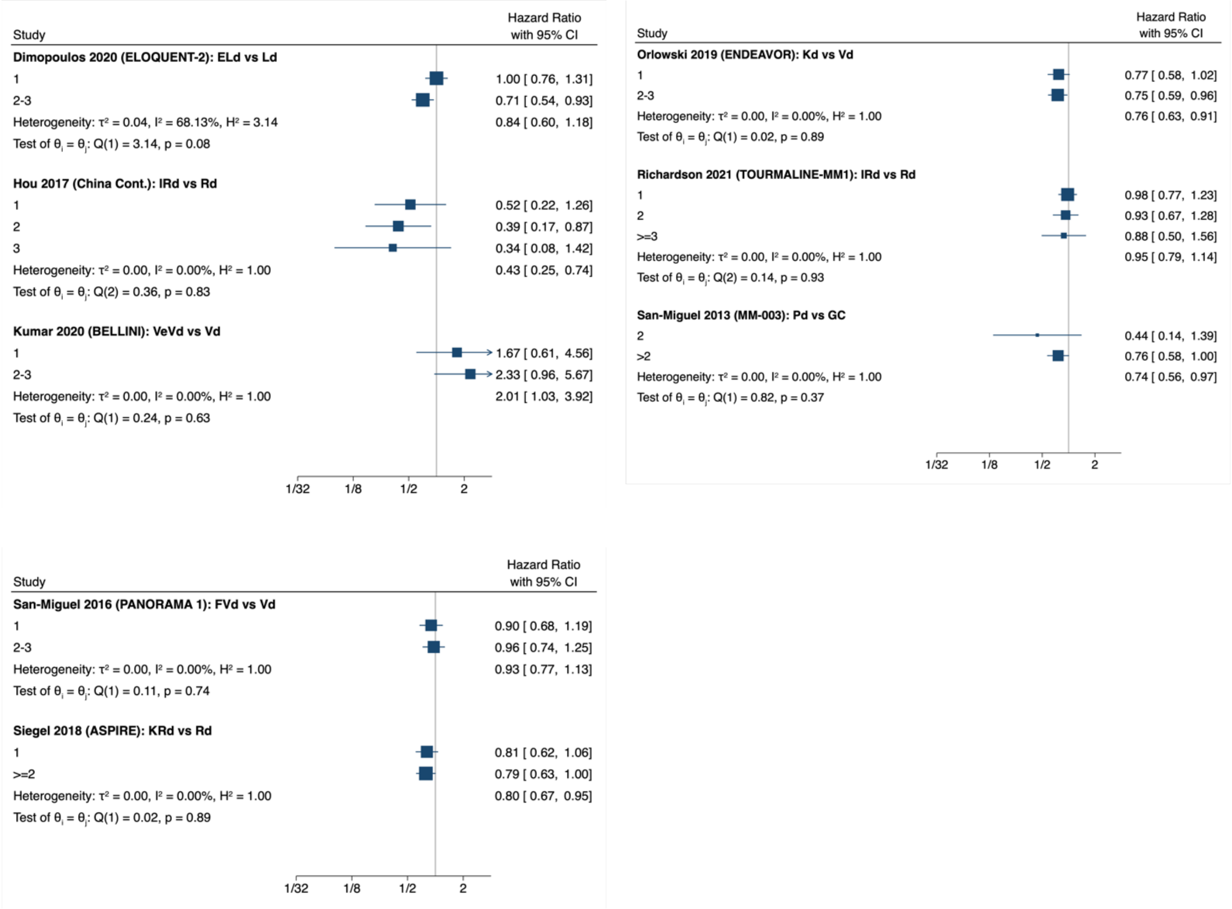

#### *Supplementary Figure 4: Estimates of ratios of hazard ratios for OS*

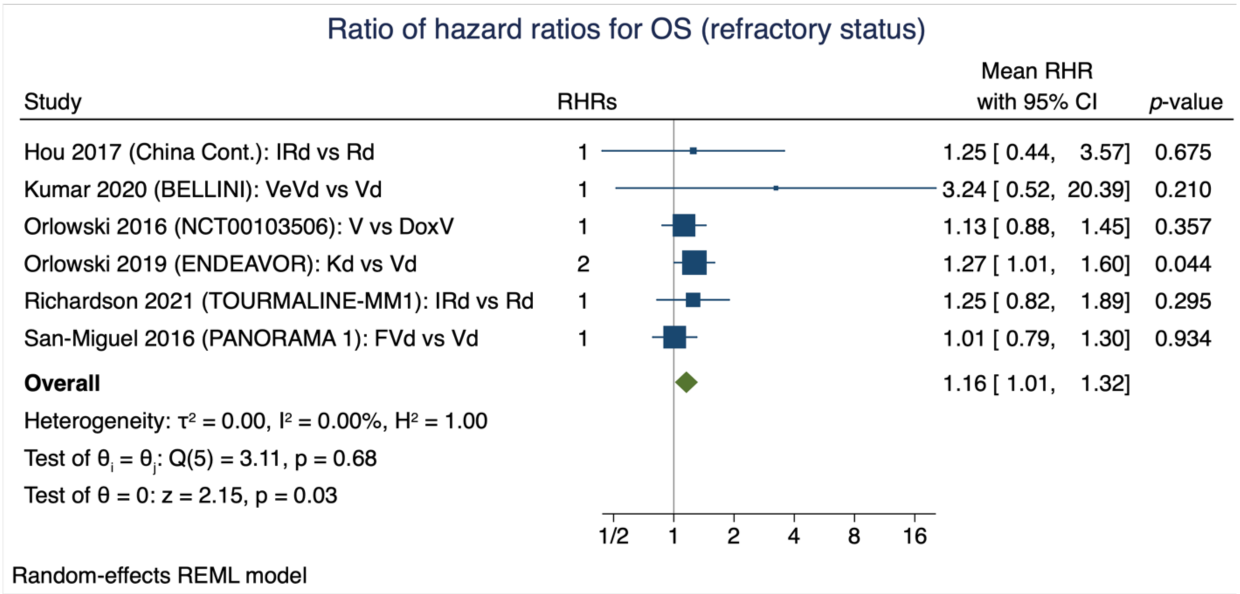

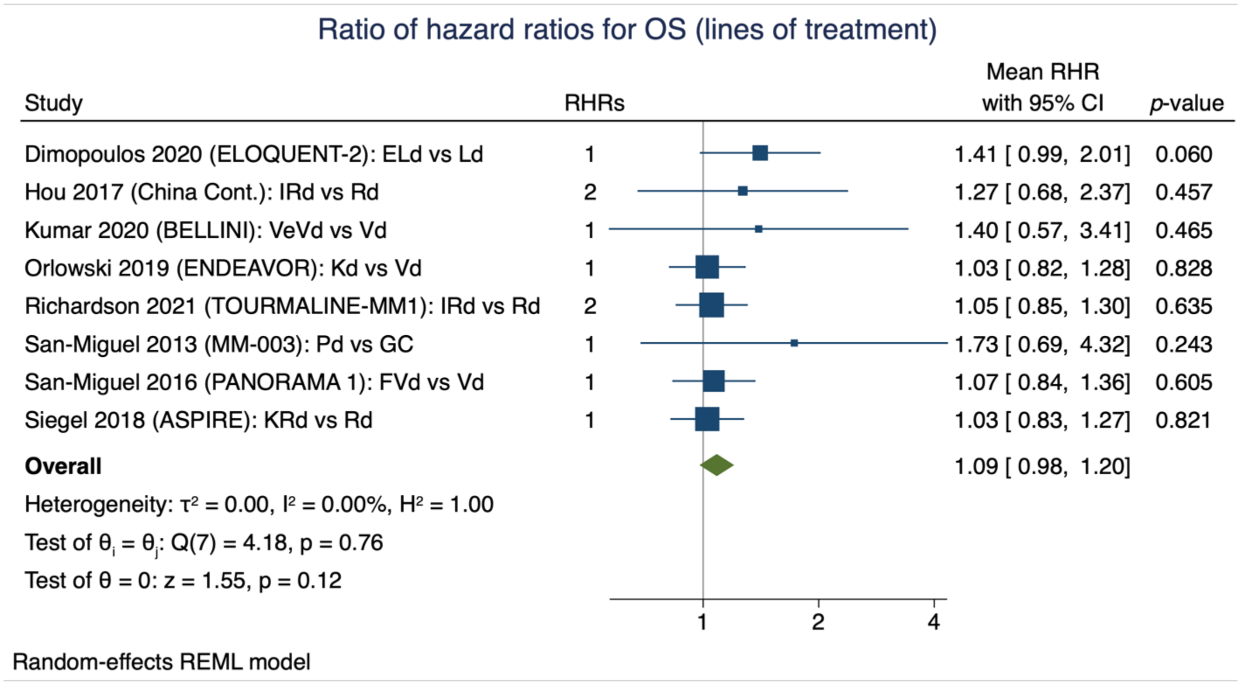

**Working Paper** — This document has not yet been submitted for peer review and may be revised.

### Supplementary Methods

#### *Search Strategy*

##### *Search strategy from February-August 2020*

Database: Ovid MEDLINE(R) and Epub Ahead of Print, In-Process & Other Non-Indexed Citations, Daily and Versions(R) <1946 to September 03, 2020>

Search Date: 07.09.2020

1 Clinical Trial, Phase III/ or exp Randomized Controlled Trial/ (515980)

2 (cross-over or crossover or ((double or single or triple) adj blind*) or (phase adj ("3" or "III")) or placebo or random*).tw,kw,kf. (1310409)

3 1 or 2 (1408374)

4 exp Multiple Myeloma/ (41318)

5 (myeloma* or Kahler disease).tw,kw,kf. (54703)

6 4 or 5 (61942)

7 Bortezomib/ (5694)

8 bortezomib.tw,kw,kf. (8199)

9 carfilzomib.tw,kw,kf. (950)

10 daratumumab.tw,kw,kf. (632)

11 elotuzumab.tw,kw,kf. (259)

12 isatuximab.tw,kw,kf. (65)

13 ixazomib.tw,kw,kf. (330)

14 Lenalidomide/ (2668)

15 lenalidomide.tw,kw,kf. (4307)

16 Panobinostat/ (513)

17 panobinostat.tw,kw,kf. (711)

18 Pomalidomide/ (0)

19 pomalidomid*.tw,kw,kf. (693)

20 Thalidomide/ (8932)

21 thalidomid*.tw,kw,kf. (8148)

22 7 or 8 or 9 or 10 or 11 or 12 or 13 or 14 or 15 or 16 or 17 or 18 or 19 or 20 or 21 (21608)

23 3 and 6 and 22 (1033)

24 (exp Animals/ or exp Animal Experimentation/) not Humans/ (4731206)

25 (animal* or dog or dogs or "in vitro" or mouse or mice or rat or rats or rodent*).ti. (1863666)

26 24 or 25 (5212771)

27 23 not 26 (1026)

28 (202002* or 202003* or 202004* or 202005* or 202006* or 202007* or 202008*).dt. (850427)

29 27 and 28 (63)

Database: Embase <1974 to 2020 September 03>

Search Date: 07.09.2020

1 Phase 3 Clinical Trial/ or exp Randomized Controlled Trial/ or Crossover Procedure/ or Double-Blind Procedure/ or Single-Blind Procedure/ (711831)

2 (cross-over or crossover or ((double or single or triple) adj blind*) or (phase adj ("3" or "III")) or placebo or random*).tw,kw. (1781327)

3 1 or 2 (1891248)

4 Multiple Myeloma/ (76323)

5 (myeloma* or Kahler disease).tw,kw. (83278)

6 4 or 5 (98992)

7 Bortezomib/ (30750)

8 bortezomib.tw,kw. (18786)

9 Carfilzomib/ (4194)

10 carfilzomib.tw,kw. (2735)

11 Daratumumab/ (2487)

12 daratumumab.tw,kw. (1762)

13 Elotuzumab/ (1077)

14 elotuzumab.tw,kw. (619)

15 Isatuximab/ (314)

16 isatuximab.tw,kw. (153)

17 Ixazomib/ (1328)

18 ixazomib.tw,kw. (841)

19 Lenalidomide/ (19234)

20 lenalidomide.tw,kw. (12112)

21 Panobinostat/ (3806)

22 panobinostat.tw,kw. (1602)

23 Pomalidomide/ (3315)

24 pomalidomid*.tw,kw. (2101)

25 Thalidomide/ (27428)

26 thalidomid*.tw,kw. (13028)

27 7 or 8 or 9 or 10 or 11 or 12 or 13 or 14 or 15 or 16 or 17 or 18 or 19 or 20 or 21 or 22 or 23 or 24 or 25 or 26 (66213)

28 3 and 6 and 27 (3613)

29 (exp Animal/ or exp Animal Experiment/) not exp Human/ (4977847)

30 (animal* or dog or dogs or "in vitro" or mouse or mice or rat or rats or rodent*).ti. (2023123)

31 29 or 30 (5420566)

32 28 not 31 (3540)

33 (202002* or 202003* or 202004* or 202005* or 202006* or 202007* or 202008*).dc. (1274593)

34 32 and 33 (173)

*Search strategy from August 2020 – March 2021*

Database: Ovid MEDLINE(R) and Epub Ahead of Print, In-Process, In-Data-Review & Other Non-Indexed Citations, Daily and Versions(R) <1946 to March 12, 2021>

Search Date: 14.03.2021

1 Clinical Trial, Phase III/ or exp Randomized Controlled Trial/ (528869)

2 (cross-over or crossover or ((double or single or triple) adj blind*) or (phase adj ("3" or "III")) or placebo or random*).tw,kw,kf. (1362145)

3 1 or 2 (1461425)

4 exp Multiple Myeloma/ and (Relaps* or Refractory).mp. [mp=title, abstract, original title, name of substance word, subject heading word, floating sub-heading word, keyword heading word, organism supplementary concept word, protocol supplementary concept word, rare disease supplementary concept word, unique identifier, synonyms] (4797)

5 ((relaps* or refractory) adj3 (myeloma* or Kahler* disease)).tw,kf,kw. (3022)

6 4 or 5 (5471)

7 Bortezomib/ (5908)

8 bortezomib.tw,kw,kf. (8517)

9 carfilzomib.tw,kw,kf. (1045)

10 daratumumab.tw,kw,kf. (788)

11 elotuzumab.tw,kw,kf. (288)

12 isatuximab.tw,kw,kf. (87)

13 ixazomib.tw,kw,kf. (369)

14 Lenalidomide/ (2811)

15 lenalidomide.tw,kw,kf. (4530)

16 Panobinostat/ (528)

17 panobinostat.tw,kw,kf. (750)

18 Pomalidomide/ (0)

19 pomalidomid*.tw,kw,kf. (759)

20 Thalidomide/ (9087)

21 thalidomid*.tw,kw,kf. (8279)

22 7 or 8 or 9 or 10 or 11 or 12 or 13 or 14 or 15 or 16 or 17 or 18 or 19 or 20 or 21 (22486)

23 3 and 6 and 22 (509)

24 (exp Animals/ or exp Animal Experimentation/) not Humans/ (4799583)

25 (animal* or dog or dogs or "in vitro" or mouse or mice or rat or rats or rodent*).ti. (1893870)

26 24 or 25 (5295514)

27 23 not 26 (507)

28 (2020083* or 202009* or 202010* or 202011* or 202012* or 202101* or 202102*).dt. (785905)

29 27 and 28 (22)

Database: Embase <1974 to 2021 March 12>

Search Date: 14.03.2021

1 Phase 3 Clinical Trial/ or exp Randomized Controlled Trial/ or Crossover Procedure/ or Double-Blind Procedure/ or Single-Blind Procedure/ (748472)

2 (cross-over or crossover or ((double or single or triple) adj blind*) or (phase adj ("3" or "III")) or placebo or random*).tw,kw. (1866956)

3 1 or 2 (1979179)

4 exp Multiple Myeloma/ and (Relaps* or Refractory).mp. [mp=title, abstract, heading word, drug trade name, original title, device manufacturer, drug manufacturer, device trade name, keyword, floating subheading word, candidate term word] (15443)

5 ((relaps* or refractory) adj3 (myeloma* or Kahler* disease)).tw,kw. (6859)

6 4 or 5 (16033)

7 Bortezomib/ (32207)

8 bortezomib.tw,kw. (19713)

9 Carfilzomib/ (4642)

10 carfilzomib.tw,kw. (2991)

11 Daratumumab/ (3057)

12 daratumumab.tw,kw. (2186)

13 Elotuzumab/ (1186)

14 elotuzumab.tw,kw. (676)

15 Isatuximab/ (387)

16 isatuximab.tw,kw. (198)

17 Ixazomib/ (1508)

18 ixazomib.tw,kw. (951)

19 Lenalidomide/ (20351)

20 lenalidomide.tw,kw. (12796)

21 Panobinostat/ (4011)

22 panobinostat.tw,kw. (1691)

23 Pomalidomide/ (3635)

24 pomalidomid*.tw,kw. (2312)

25 Thalidomide/ (28149)

26 thalidomid*.tw,kw. (13408)

27 7 or 8 or 9 or 10 or 11 or 12 or 13 or 14 or 15 or 16 or 17 or 18 or 19 or 20 or 21 or 22 or 23 or 24 or 25 or 26 (69342)

28 3 and 6 and 27 (1874)

29 (exp Animal/ or exp Animal Experiment/) not exp Human/ (5079959)

30 (animal* or dog or dogs or "in vitro" or mouse or mice or rat or rats or rodent*).ti. (2066725)

31 29 or 30 (5536425)

32 28 not 31 (1841)

33 (2020083* or 202009* or 202010* or 202011* or 202012* or 202101* or 202102*).dc. (1351126)

34 32 and 33 (62)

35 limit 34 to embase status (26)

*Search strategy for ongoing studies*

Search date: June 2021

Search line: Multiple Myeloma AND (Relapse OR Refractory)

#### *Missing data*

We imputed standard errors from confidence intervals using standard Cochrane methodology where necessary. We did not attempt to impute missing estimates (e.g., from Kaplan-Meier plots or median survival times) as this would have been excessively time-consuming and inaccurate. We did not contact study authors to request missing stratified estimates.

#### *Further details on the simulation study*

With respect to refractory status, a dichotomous variable, we assumed that effect modification could apply to between 0% and 100% of patients within a trial (this was modelled this using a uniform distribution). Because RHR is defined in a way that discards direction of effect modification, we ensured that direction was consistent within treatment comparison, but could vary between comparisons (direction of effect within comparison was modelled using a uniform Bernoulli distribution).

With respect to LOT, a categorical variable with up to four levels in the real PFS data (e.g., 1, 2, 3, and >3 LOT), we modelled a worse-case scenario by assuming the variable has four levels in all simulated studies, that effect modification consistently increases or decreases with LOT (e.g., that HRs are larger in 3 versus 2 LOT, and larger in 2 versus 1 LOT), and that modification "compounds" over LOT categories (analogous to how interest on savings compounds over investment time), consistent with how RHR is defined.

It may appear necessary to simulate the “total” impact of both refractory status and LOT. However, this is unrealistic because these variables do not modify effect independently: to a large extent, we expect that the RHR for LOT already accounts for effect modification due to refractory status. In other words, most patients who have received two previous lines of treatment will presumably also be refractory to those two treatments. (Considering our broad definition of refractory status leads to a slightly more complex argument, but to the same conclusion.) We therefore did not simulate a combination of the impact of both refractory status and LOT, as to do so would “double count” any effect modification and, at best, provide a presumably quite large and uninformative upper bound on the percentage of estimates that would be expected to differ.

***Ratio of hazard ratios and its sampling variance***

Equation (*1*) defines ratio of hazard ratio (RHR) on the logarithmic scale.

$$\log \hat{R}_{i,j} = \left|\log \hat{H}_{i,j} - \log \hat{H}_{i,j-1}\right|, \ \ \forall\, i \geq 1, j > 1, \tag{1}$$

where $\hat{H}_{i,j}$ is the hazard ratio reported for stratum *j* of trial *i*; studies and strata are indexed from one; and |•| denotes absolute value. To facilitate inverse variance-weighted meta-analysis, we seek the sampling variance on log RHR, $\log \hat{R}_{i,j}$. For brevity, we will neglect trial indices and consider two strata, *i* = 1 and *i* = 2, whose stratified hazard ratios may be correlated. Recall that $\mathrm{Var}(X) = \mathrm{E}[X^2] - \mathrm{E}^2[X]$. This gives:

$$\begin{aligned} \mathrm{Var}(|\log H_1 - \log H_2|) &= \mathrm{E}[|\log H_1 - \log H_2|^2] - \mathrm{E}^2[|\log H_1 - \log H_2|] \\ &= \mathrm{E}[(\log H_1 - \log H_2)^2] - \mathrm{E}^2[|\log H_1 - \log H_2|] \end{aligned} \tag{2}$$

Let

$$\boldsymbol{X} = \begin{bmatrix} \log H_1 \\ \log H_2 \end{bmatrix} \sim N\left(\begin{bmatrix} \log \hat{H}_1 \\ \log \hat{H}_2 \end{bmatrix}, \begin{bmatrix} v_1 & \rho\sqrt{v_1}\sqrt{v_2} \\ \rho\sqrt{v_1}\sqrt{v_2} & v_2 \end{bmatrix}\right), \tag{3}$$

model the joint uncertainty on the log HRs, where $v_i$ is the sampling variance for the *i*-th stratum and $\rho$ is the correlation between HRs. Expected value $\mathrm{E}[(\log H_1 - \log H_2)^2]$ can be obtained via the transform $[1 \ \ -1]\boldsymbol{X}$, which yields a univariate random variable with mean $\theta = \log \hat{H}_1 - \log \hat{H}_2$ and variance $\varphi^2 = v_1 + v_2 - 2\rho\sqrt{v_1}\sqrt{v_2}$, which gives $\mathrm{E}[(\log H_1 - \log H_2)^2] = \theta^2 + \varphi^2$. Squared expected value $\mathrm{E}^2[|\log H_1 - \log H_2|]$ can be shown to be $\frac{1}{\pi}e^{-\theta^2/\varphi^2}\left(\sqrt{2}\varphi+\sqrt{\pi}e^{\theta^2/\varphi^2}\theta\,\mathrm{erf}\frac{\theta}{\sqrt{2}\varphi}\right)^2$. The sampling variance of $|\log H_1-\log H_2|$ is therefore

$$\mathrm{Var}(|\log H_1-\log H_2|)=\theta^2+\varphi^2-\frac{1}{\pi}e^{-\theta^2/\varphi^2}\left(\sqrt{2}\varphi+\sqrt{\pi}e^{\theta^2/\varphi^2}\theta\,\mathrm{erf}\frac{\theta}{\sqrt{2}\varphi}\right)^2, \tag{4}$$

where erf is the error function. Because correlation $\rho$ is unknown, we choose $\rho=0$, which maximally favors the effect modification hypothesis in the sense that knowing a HR for one stratum provides no information about the other.

**Appendix: Conditional *P*-scores for treatment ranking when some treatments are not competitors**

***Introduction***

A key result of a network meta-analysis (NMA) is a ranking of treatments, with respect to a specific outcome, from best to worst. It is not sensible to rank treatments by point estimate, because point estimates lack precision due to sampling error and possibly other factors; further, precisions will vary due to differences in trial sample sizes and the network topology. A treatment ranking assumes that all treatments are competitors (scientifically, with respect to a specific outcome, rather than in terms of another competitive arena such as regulatory approval or price). This is not always the case in relapsed refractory multiple myeloma (RRMM) because patients can be refractory to specific treatments. Refractoriness to a specific treatment or treatment component is a *de facto* treatment modifier of effect estimates involving that treatment or component. The quantities by which treatments are ranked (see below) must therefore be conditioned on the set of treatments that are competitors. This appendix proposes a method called conditional *P*-scores — a simple modification of the original method — that addresses this problem.

#### *Theory*

More formally, treatment ranks are ordinal values that are computed from continuous values that assess the extent of evidence that each treatment is *superior* (i.e., better than all the other treatments). Two methods have gained widespread use in the NMA literature: Surface Under the Cumulative RAnking (SUCRA) curve scores[1], which are applicable to Bayesian NMAs; and *P*-scores (cf. *p*-values)[2], which are frequentist equivalents to SUCRA scores.

If each trial included in an NMA recruited *only*[*] patients not refractory to the treatments being compared in the trial — it is difficult to imagine a trial that does the opposite receiving ethical approval — then the network meta-analytical effect estimates are conditioned on patients not being refractory to any of the treatments included in the NMA. The NMA results may then be used to rank treatments, but only for non-refractory patients. To rank treatments for patients who are refractory to specific treatments, it is necessary to compute SUCRA values or *P*-scores conditionally with respect to the treatments to which patients are refractory.

[*] I.e., an overwhelming majority of patients, such that the trial would be "fair"; we are aware of some trials that recruited small proportions of patients who were refractory to specific components included in the treatments being compared.

*P*-scores are computed from all pairs of effect estimates — i.e., the matrix of point estimates, ***B*** (with elements $b_{i,j}$, where *i* and *j* index treatment), and the associated matrix of variances, ***V*** (with elements $v_{i,j}$). The following presentation is a slightly simplified version of that of Rücker and Schwarzer[2]. The vector of *P*-scores, ***p***, is given by

$$\boldsymbol{p} = \frac{\mathbf{1}}{\mathbf{1}-\boldsymbol{k}}(\boldsymbol{P}\ \mathbf{1}_{\boldsymbol{k}} - \mathrm{diag}\,\boldsymbol{P}) \tag{5}$$

where: there are *k* treatments; $\mathbf{1}_k$ is a $k \times 1$ vector of ones; the elements of ***P*** are

$$P_{i,j} = \begin{cases} 1 - \Phi\left(\left|\frac{b_i - b_j}{\sqrt{v_{i,i}+v_{j,j}}}\right|\right) & \text{if } b_i \le b_j \\ \Phi\left(\left|\frac{b_i - b_j}{\sqrt{v_{i,i}+v_{j,j}}}\right|\right) & \text{otherwise} \end{cases}; \tag{6}$$

and ϕ is the cumulative distribution function for the standard normal.

For patients who are refractory to a specific set of *r* treatments, conditional *P*-scores can be computed for the remaining (*k* - *r*) treatments to which the patients are not refractory as follows. First, form the (non-leading) principal submatrices ***B**** and ***V**** by removing from ***B*** and ***V*** the rows and columns that correspond to the *r* treatments. Then use ***B**** and ***V**** in place of ***B*** and ***V*** to compute *P*-scores for the remaining (*k* - *r*) treatments.

#### ***References***

1. Salanti G, Ades AE, Ioannidis JP. Graphical methods and numerical summaries for presenting results from multiple-treatment meta-analysis: an overview and tutorial. *J Clin Epidemiol*. 2011; 64(2):163–71.

2. Rücker G, Schwarzer G. Ranking treatments in frequentist network meta-analysis works without resampling methods. *BMC Med Res Methodol*. 2015; 15(58).
